# Modelling suggests blood group incompatibility may substantially reduce SARS-CoV-2 transmission

**DOI:** 10.1101/2020.07.13.20152637

**Authors:** Peter J. I. Ellis

## Abstract

Several independent datasets suggest blood type A is over-represented and type O under-represented among COVID-19 patients. Here, I model a scenario in which ABO transfusion incompatibility reduces the chance of a patient transmitting the virus to an incompatible recipient. Comparison of model outputs to published data on COVID-19 prevalence indicates that if this scenario holds true, ABO incompatibility may reduce virus transmissibility by 60% or more. Paradoxically, however, targeted vaccination of either high-susceptibility type A or “super-spreader” type O individuals is less effective than random vaccination at blocking community spread of the virus. Instead, the key is to maintain blood type diversity amongst the remaining susceptible individuals. I stress that these results illustrate a theoretical model of ABO blood group interaction with virus transmission and require confirmation by observation.

## Introduction

Several recent published studies and preprints have suggested that the prevalence of COVID-19 disease varies by blood type, with type A being relatively susceptible and type O being less susceptible [1-5]. Puzzlingly, however, there is no difference in the severity of disease, with the case fatality ratio (CFR) and the probability of progressing to intensive care appearing independent of blood type. This discrepancy between incidence and severity data has led some authors to challenge the aforementioned findings [6]. Although not remarked on to date, in the majority of these studies type AB appears even more susceptible than type A. Thus, the relative risk of infection is AB > A > B > O, with type A and type B alleles functioning codominantly to increase risk. This is immediately reminiscent of the rules governing blood transfusion compatibility.

Here, I investigate the behaviour of “ABO-interference”: a model of epidemic spread in which the transmission of severe acute respiratory syndrome coronavirus 2 (SARS-CoV-2, the causal agent of COVID-19) is dependent on the ABO blood type compatibility between an infected individual and the susceptible people they encounter. Mechanistically, this models a scenario in which infectious virions acquire the glycosylation pattern and hence the ABO antigen status of their current host. This in turn allows shed virions to be “rejected” by incompatible recipients, blocking the initial infective step. This immediately explains the lack of correlation with disease severity, since once an infection is established, virions produced within the new host are necessarily self-compatible and able to spread freely between cells.

The plausibility of this hypothesis has already been established by work on HIV [7-8], and was also previously proposed for the 2003 epidemic of SARS [9-10], for measles [11], and indeed for enveloped viruses in general [12-13]. SARS-CoV-2 has an outer lipid membrane containing spike, membrane and envelope (S, M and E) proteins, all of which are exposed to immune recognition and any or all of which may be glycosylated. Structural studies show that S is heavily glycosylated, including fucosylated glycans that may potentially bear ABO determinants [14-15]. The glycosylation status of the M and E proteins has not yet been characterised, nor the glycosylation status of the membrane lipids. Experimental work using pseudotyped virus suggests that transmission of the closely related SARS virus can be blocked by anti-A antibodies when virus particles are grown in an A-expressing cell line [10].

This study also showed via modelling that ABO-interference can reduce the progress of an epidemic dependent on the magnitude of the block to transmission and the local population structure. However, despite the mechanistic plausibility of this hypothesis and the preliminary data from the SARS epidemic, there has as yet been no detailed modelling exploring the implications of ABO-interference for the relative susceptibility of individuals with different blood types at different stages of the epidemic, or for vaccination strategies. In this analysis, I develop an extended SIR (<u>**S**</u>usceptible, <u>**I**</u>nfected, <u>**R**</u>ecovered) epidemiological model which allows for a partial or total block to virus transmission from an infected patient to an incompatible recipient, and explore the implications of this model for epidemic progression and for vaccination strategies.

## Results

### Modelling ABO-interference with virus transmission

The simplest of all epidemic models assumes a homogeneously mixing population divided into susceptible, infectious and recovered groups or “compartments”. This yields three variables S, I and R, which respectively represent the proportion of the population with each status. Such a model is completely described by two parameters *β* and *ν*, representing the rate constants for infection and recovery respectively. The duration of the infectious period *d* is given by 1 / *ν*. All simulations presented here are based on *d* =7 days, and none of the results presented are sensitive to this parameter. R0 is given by the product *β*.*ν* and represents the basic reproduction number, i.e. the mean number of individuals infected by a typical infectious individual during their illness, in the context of a completely susceptible population [16]. To this model I add a further parameter *ρ* where 0 ≤ *ρ* < 100%, representing the relative probability of cross-type infection (see Methods for full details). In this extended model, R0 is not a well-defined quantity since even in a fully susceptible population the current effective R value, denoted R(t), depends on *ρ* and on the blood type distributions in both S and I. Below, I use R_max_ to indicate the maximal possible value of R(t). This will be observed when the population is fully susceptible and transmission is unimpeded, i.e. when *ρ* = 100% or all currently infected individuals are type O.

### Type O individuals are critical for epidemic progression when *ρ* is low or zero

**Figure 1 A-E** illustrates the effect of complete ABO incompatibility on transmission dynamics in a typical Western European background with a population blood type distribution of 38% A / 14% B / 4% AB / 44% O, R_max_ = 4 and *ρ* = 0. If the index case is type A, an epidemic occurs within the A population, and also propagates to AB (since A→AB is a permissible vector of transmission). If the index case is type B or AB, the epidemic never becomes established since there are too few compatible susceptible individuals to sustain transmission. If the index case is type O, epidemics occur within all four populations. In all cases the progress of the epidemic is profoundly suppressed relative to an epidemic with no ABO-interference, however the epidemic stemming from a type O index case is four times the size of that stemming from a type A index case.

**Figure 1.**
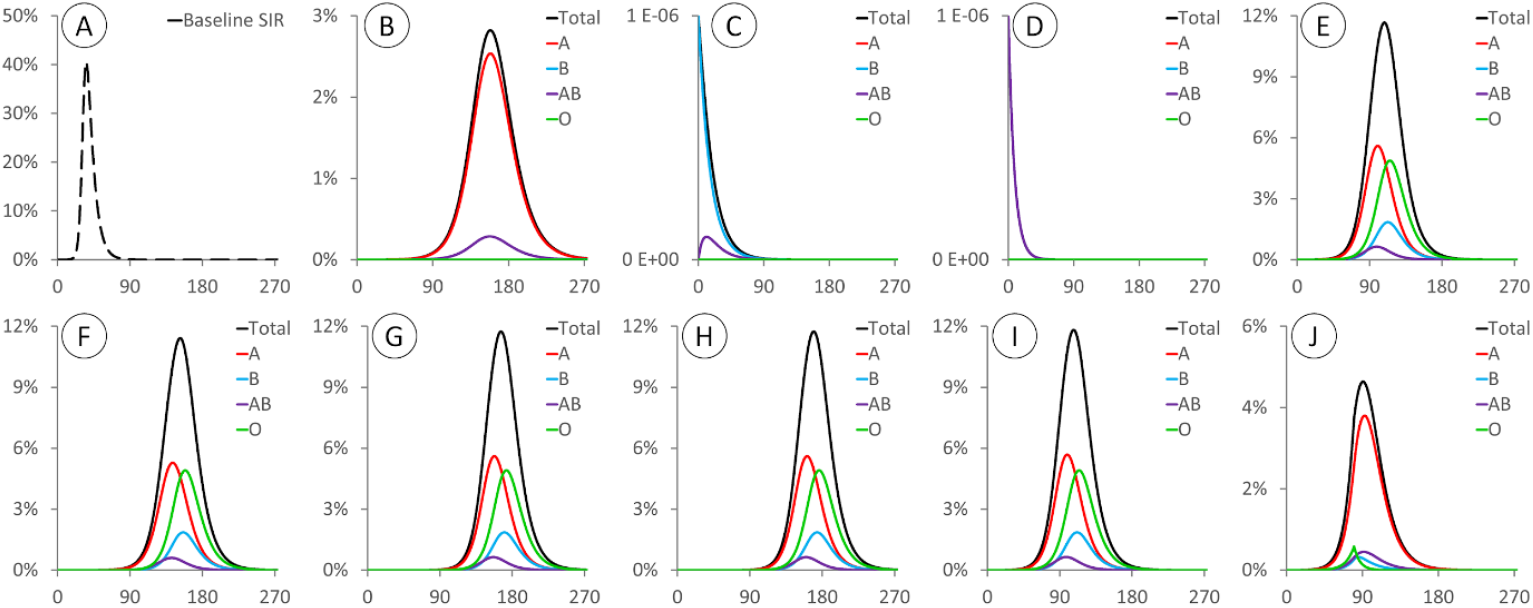
X axis: days. Y axis: proportion of the population infected (different scales used in each panel to show detail). All epidemics are based on a typical “Western” blood type distribution of 38% A / 14% B / 4% / AB / 44% O and initialised at t=0 with 1/1,000,000 of the population infected. **A:** Epidemic with R_max_ = 4 and no ABO-interference with transmission, peaking at 40.5% of the population on day 35. **B/C/D/E:** Epidemics with R_max_ = 4 and *ρ* = 0, with index cases of type A/B/AB/O respectively. **F/G/H/I:** Epidemics with R_max_ = 4 and *ρ* = 0.1%, with index cases of type A/B/AB/O respectively. **J:** Epidemics with R_max_ = 4 and *ρ* = 0.1%, with index cases of type B and all susceptible type O individuals assumed to be vaccinated at day 80.

Importantly, if *ρ* > 0 and there is *<u>any</u>* degree of spread between incompatible blood types, at least some proportion of the O population will become infected. Thereafter the epidemic will behave similarly to one seeded by type O index cases, as shown in **Figure 1 F-I** for *ρ* = 0.1%. If the index case is non-O [figure **1F/G/H**], this delays the onset of the epidemic and the date of the peak relative to a type O index case [figure **1I**], but affects neither the size of the peak nor how it progresses between and within different population compartments. For *ρ* = 0.1%, the epidemic among B individuals is not sustainable from A→B cross-transmission alone but is dependent on the epidemic among O individuals. If these are removed from the susceptible population after 80 days (e.g. by vaccination), this immediately halts the spread among B individuals [**Figure 1J**]. The epidemics in A and AB are also affected, but to a more modest degree since the epidemic is self-sustaining amongst A individuals and these can then transmit to AB.

### A quasi steady state is obtained when all blood types are infected

Whenever infection is present among individuals with all four blood types (i.e. there is at least one type O index case, or *ρ* > 0), the blood type distribution among infected people rapidly converges upon a new equilibrium that is significantly skewed relative to the initial population distribution. This equilibration process causes R(t) to also settle on an equilibrium value during the early stages of the epidemic, hereafter called R_steady_. This is a rapid process, with R(t) reaching R_steady_ within a few serial intervals: in these simulations this equates to ∼0.1% of the population being infected. Therefore, in real-world epidemics subject to ABO-interference, estimates of R0 based on population statistics are likely to actually measure R_steady_ and thus underestimate the true value of R_max_. **Figure 2** illustrates the nature of this steady state, which depends on *ρ* (compare rows **A,B,C**), but is independent of R_max_ (compare rows **C,D,E**) and the number and blood type of the index cases (not shown). R_steady_ also depends on the background population blood type distribution (compare rows **C,F,G**). Importantly, during this steady state, R_steady_ is a consistent multiple of R_max_, indicating that ABO-interference suppressed the epidemic with equal efficiency regardless of the underlying infectiousness of the pathogen. Similarly, the relative risk to each blood type is also dependent only on *ρ* and the background population blood type distribution. Thus, regardless of whether the epidemic in any given region is progressing quickly or slowly, the relative risk to each blood type will be predictable from the population blood group frequencies and the specific value of *ρ*.

**Figure 2.**
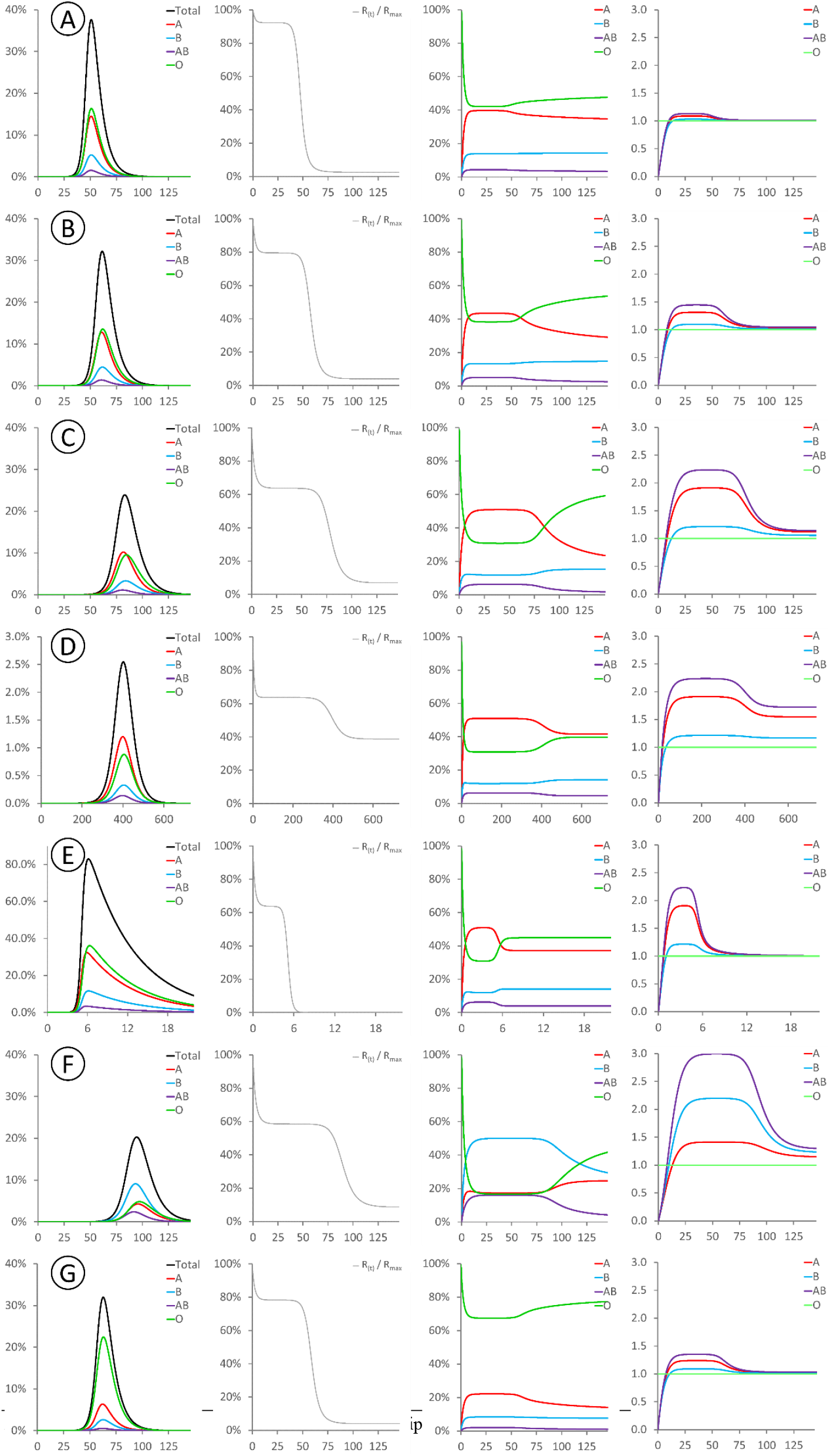
These panels show the evolution of various modelled parameters for epidemics initialised under varying conditions. All epidemics were seeded at with type O index cases, and thus R(t) = R_max_ at time t=0. For all sub-panels, X axis denotes days. Y axes: (first column) proportion of the population infected; (second column) R(t) as a fraction of R_max_; (third column) distribution of blood types among currently infected individuals; (fourth column) cumulative risk of infection for each blood type relative to type O. Rows **A-E** are initialised with the same typical Western population blood type distribution used in Figure 1. **A:** R_max_ = 4, *ρ* = 80%. **B:** R_max_ = 4, *ρ* = 50%. **C:** R_max_ = 4, *ρ* = 20%. **D:** R_max_ = 2, *ρ* = 20%. **E:** R_max_ = 40, *ρ* = 20%. Row **F:** R_max_ = 4, *ρ* = 20%, initialised with an Indian population blood type distribution of 21.4%/39.9%/9.4%/29.3% type A/B/AB/O [17]. Row **G:** R_max_ = 4, *ρ* = 20%, initialised with a Peruvian population blood type distribution of 18.9%/8.1%/1.6%/71.4% type A/B/AB/O [18].

### Estimating *ρ* for SARS-CoV-2

Since the worldwide SARS-CoV-2 pandemic appears to be still in its infancy, with low seroprevalence in all countries studied, the pandemic appears to fall within the steady-state region of the curve. **Table 1** shows data compiled from all published studies that have reported ABO frequencies amongst infected SARS-CoV-2 patients and controls [1,2,4]. These collectively cover epidemics in two regions of China, three regions of Spain, and one region each of Italy and the United States. A UK Biobank preliminary study has also reported an increased risk for type A individuals [3], but did not infer type AB frequency and was thus excluded from my analysis. For each data set, I used the control blood type distribution to predict the expected steady state distribution among infected patients for different values of *ρ*, and calculated the root mean square difference between the predictions and the observed case frequencies. Minimising this difference indicates the most likely value of *ρ*. Notably, for all data sets this lies between 25% and 40% despite the different underlying blood type frequencies and relative risk ratios in each country. For the Italian and Spanish study, data was also provided for area-matched control data from blood donors [4]: using these instead of the internal control data does not affect the result.

**Table 1.**
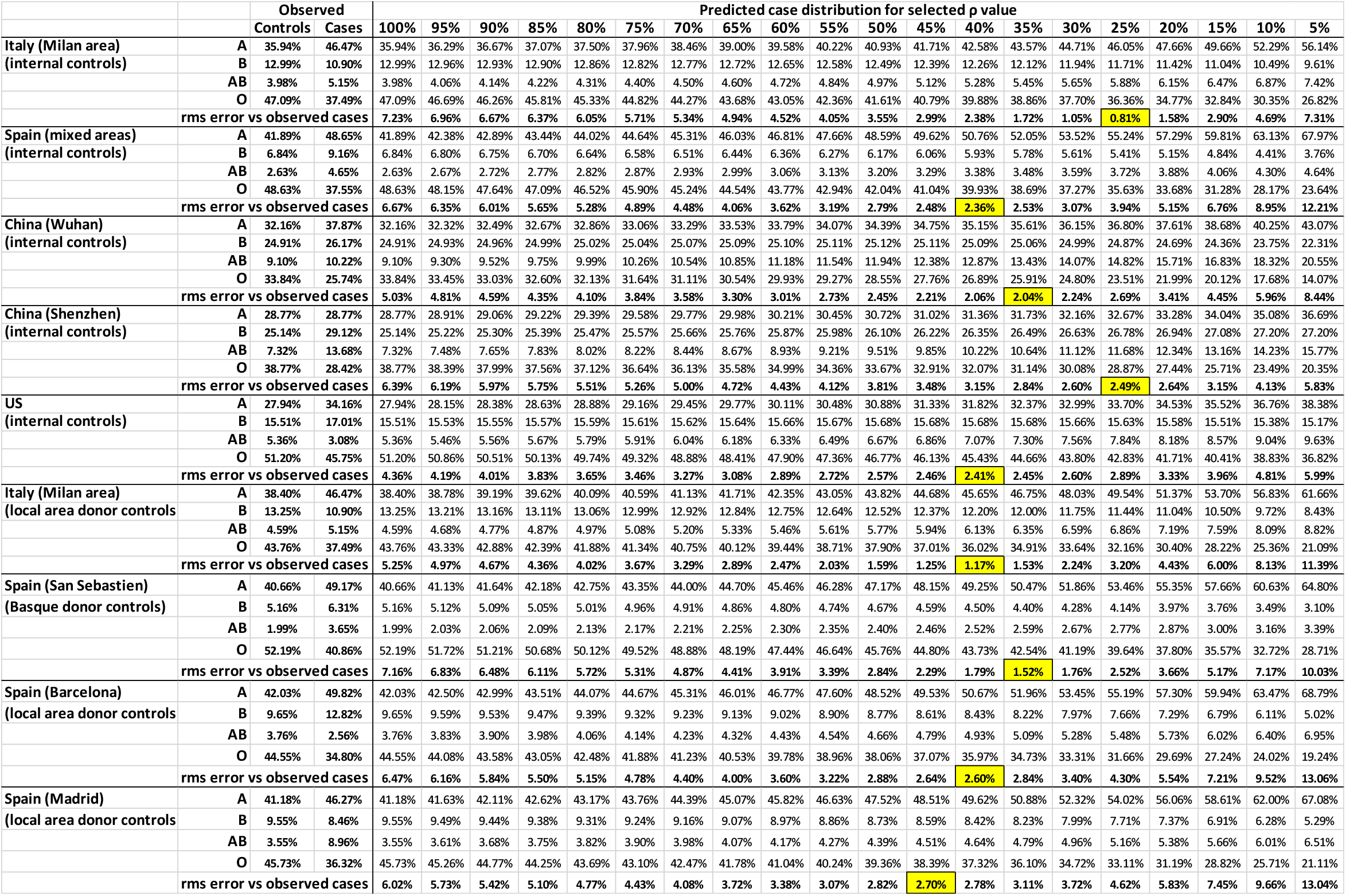
Observed case and control numbers are taken from references [1,2,4]. The Italian and Spanish data sets provided both internal and external controls. For each area, the control blood group frequencies were used to project the steady-state blood group frequencies among infected patient for varying values of *ρ*. Calculating the root mean square (rms) difference between the predicted frequencies and the actual case frequencies allows region-specific estimation of *ρ* (yellow highlight).

Three caveats apply to this analysis. Firstly, while overall prevalence in each country remains low, the local prevalence in “hotspots” is much higher [19], and infected individuals will by definition originate preferentially from these hotspots. In these areas, the epidemic may have progressed beyond the steady state region of the epidemic curve, reducing the degree of blood type skewing among the infected population and partially masking the effects of ABO-interference on virus transmission. In this case, in order to generate the observed changes in blood type frequency, then the degree of ABO-interference must be even more pronounced. i.e. the estimate of 40% is a maximum bound, and the true frequency of cross-type transmission may be even lower. Secondly, the SIR model assumes that population mixing and opportunities for transmission are independent of blood type. This is unlikely to be the case since blood relatives living together are both more likely to infect each other and more likely to share a blood type. This effect will also tend to mask the effect of ABO-interference. Thirdly: when there is nosocomial spread within a hospital, an increase in frequency of one blood type among infected patients will lead to a decrease in frequency of that blood type in the remaining uninfected hospital patients. This will exaggerate the effects of ABO-interference if uninfected hospitalised patients are used as controls.

Overall, the third of these appears not to be an important factor since for both the Italian and Spanish populations similar results are obtained using blood donor controls. Since the first two effects both bias the estimate of *ρ* upwards, the central estimate of 35-40% thus represents a best-guess upper bound for *ρ*. Intriguingly, previous data from direct contact tracing [10] imply *ρ*=47.7% for the 2003 hospital outbreak of SARS in Hong Kong, suggesting that values around this range may be a feature of coronavirus infections in general.

### Is targeting by blood type a useful vaccination strategy?

Initial preprints noting the increased risk to type A individuals have proposed that these may require additional surveillance and priority for protection. However, ABO-interference with virus transmission presents a unique and striking scenario that has not previously been modelled in detail, in which those most prone to infection are those least likely to pass it on, and vice versa. This raises the question as to whether it is more important to vaccinate the most susceptible individuals, or the most infectious individuals. **Figure 3A** shows 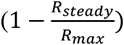, i.e. the degree to which R0 is suppressed by ABO-interference, across the full spectrum of potential ABO allele frequencies for *ρ* = 30%. ABO-interference suppresses transmission most efficiently when the allele frequency ratio is approximately 40% O / 30% A / 30% B alleles. Translating allele frequencies to blood group frequencies yields **Figure 3B**. ABO-interference suppresses transmission most efficiently when type O individuals make up 15% of the population and type A / type B individuals are present in equal proportions. A similar shape heat map is obtained for values of *ρ* = 20% and *ρ* = 90% (not shown). Vaccinating type O individuals moves the population upwards in **Figure 3B**, while vaccinating type A or B moves the population right or left respectively.

**Figure 3.**
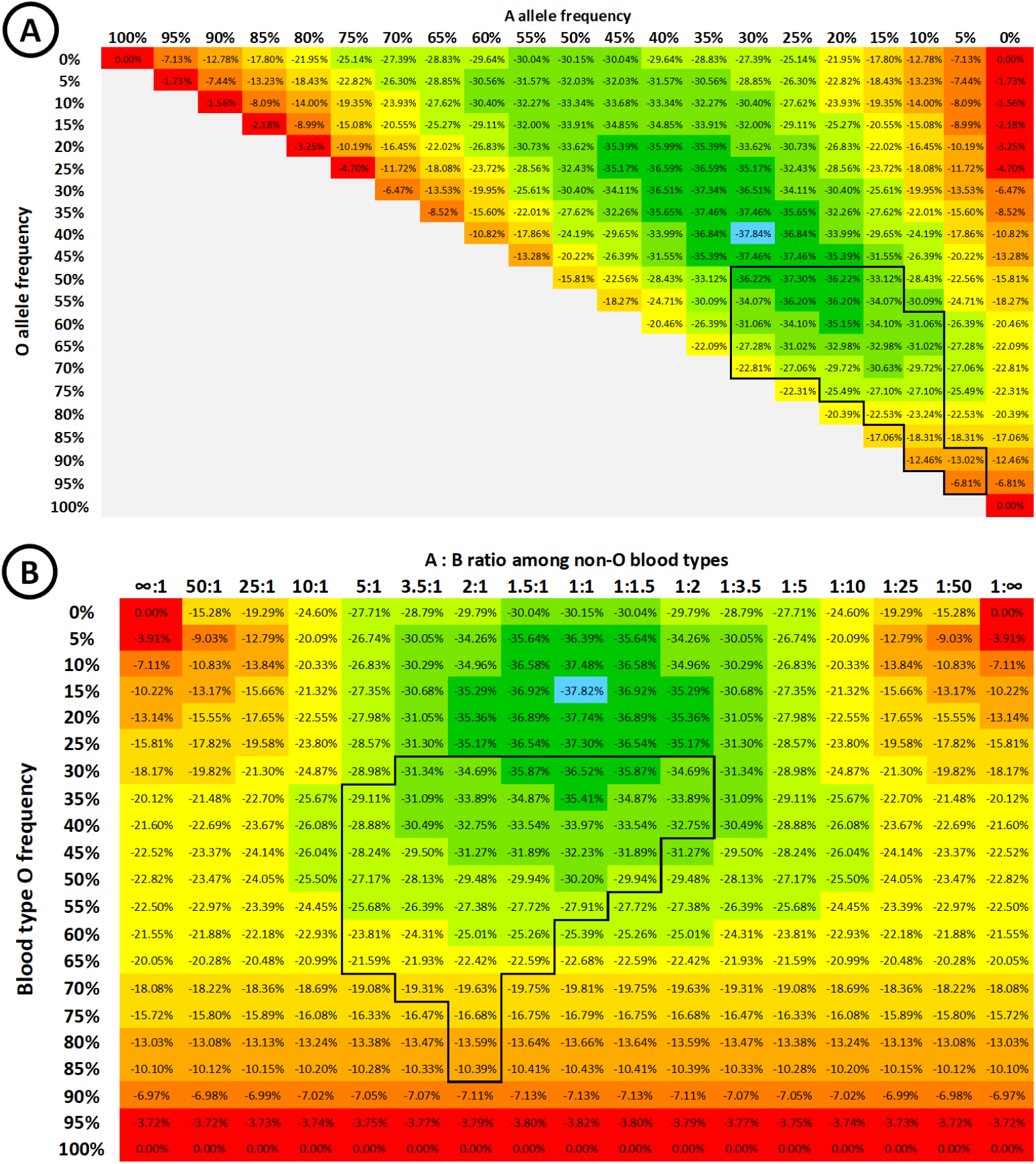
**A:** Heat map showing 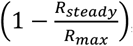, i.e. the degree of suppression of R0 during the early “steady state” portion of the epidemic, according to the population ABO allele frequencies. B allele frequency = (100% - O% - A%). **B:** Heat map showing the degree of suppression of R0 during the early stages of the epidemic, according to the background population blood type frequencies. In both heat maps, ABO allele frequencies are assumed to be in Hardy-Weinberg equilibrium. The colour gradient from red to green indicates the degree of suppression in 5% increments, with the blue highlight indicating maximal suppression of virus transmission. The boxed areas of the plots indicate allele / blood type frequencies that are typically observed in human populations.

In principle, an optimal vaccination strategy will cause the distribution among susceptible individuals to move “down” the gradient, i.e. towards more effective suppression of the epidemic. Conversely, the vaccination strategy must also be careful *<u>not</u>* to disrupt the intrinsic protection afforded by ABO-interference. To illustrate this, consider a population with 50% type A and 50% type O individuals, similar to Māori and some Polynesian populations where the type B frequency is very low [20]. In general, the predicted herd immunity threshold is 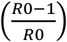, so in the absence of ABO-interference the threshold for an epidemic with an R_max_ of 3 is 66.7%. In such a population, if *ρ* = 30% then R_steady_ = 2.32, the risk for type A individuals is 1.82 times higher than type O individuals, and type O individuals are 1.54 times as infectious as type A individuals.

A well-intentioned strategy to reduce infection might prioritise vaccinating type O super-spreaders before type A. However, once all type O individuals have been immunised, the protective effect of ABO-interference is abolished since the remaining susceptible population is now exclusively type A, and an infected type A individual can freely transmit to any remaining susceptible individual. Herd immunity will therefore only be attained when the full 66.7% of the population is vaccinated. The same applies in reverse if the more vulnerable type A individuals are instead prioritised for vaccination. However, vaccinating both blood types equally produces herd immunity when 56.9% of the population is vaccinated, consistent with R_steady_ in this population. This effect is further magnified if transmission is brought down by other means, for example non-pharmaceutical interventions including social distancing. For the same population and same value of *ρ* = 30%, if R_max_ = 2 then R_steady_ = 1.55. In this case the herd immunity threshold is 50% of the population if preferentially vaccinating either type O or type A, but only 35.4% if vaccinating individuals at random [**Figure 4**].

**Figure 4.**
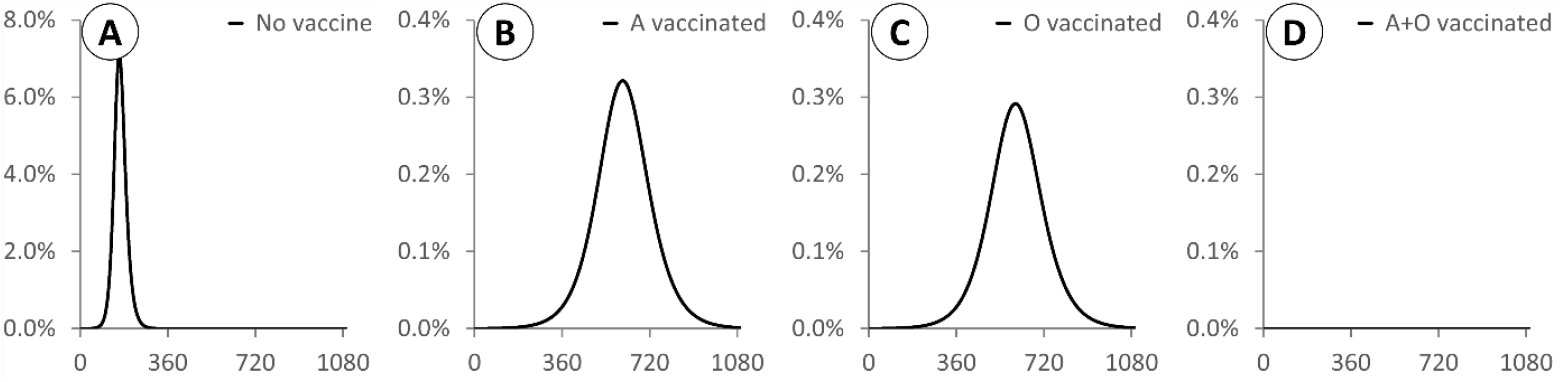
X axis: days. Y axis: proportion of the population infected (note different scale used for panel A). All epidemics are based on a population with 50% A / 50% O individuals and initialised at t=0 with type O index cases and 1/1,000,000 of the population infected. R_max_ =2 and *ρ* = 30% for all epidemics. **A:** No vaccination. **B:** 35.4% of the population vaccinated at t=0, all type A. **C:** 35.4% of the population vaccinated at t=0, all type O. **D:** 35.4% of the population vaccinated at t=0, half type A and half type O. Vaccinating O is slightly more effective than vaccinating A, but vaccinating both types leads to herd immunity at a lower threshold.

### The effect of waning immunity for a virus subject to ABO-interference

At present is it unknown whether SARS-CoV-2 leads to long-lasting immunity. The four other endemic human coronaviruses do not confer lasting immunity, and repeat infection is common. In an SIR model, when immunity is allowed to wane over time, then population spread of the virus can resume once the population level of immunity falls below the herd immunity threshold. Over time this leads to damped oscillatory behaviour, with recurrent pulses of infection converging on a final steady state. In this final steady state, by definition R(t) = 1, i.e. there is steady sustained transmission with overall infection levels neither growing nor shrinking [**Figure 5]**. The final population disease burden depends on the duration of immunity, with a shorter immune duration leading to a higher population-wide average prevalence. In the extended ABO-interference model, the relative risk to non-O blood groups is less pronounced during this final steady state compared to the initial quasi-steady state during the early phase of pandemic spread, but remains non-zero. In the final steady state, with waning immunity and recurrent infection, the relative risk to each blood type should be interpreted as a difference in the frequency of infection: e.g. if type A has a relative risk of 1.1 compared to type O, it means type A individuals are 10% more likely to suffer an infection within any given time period.

**Figure 5.**
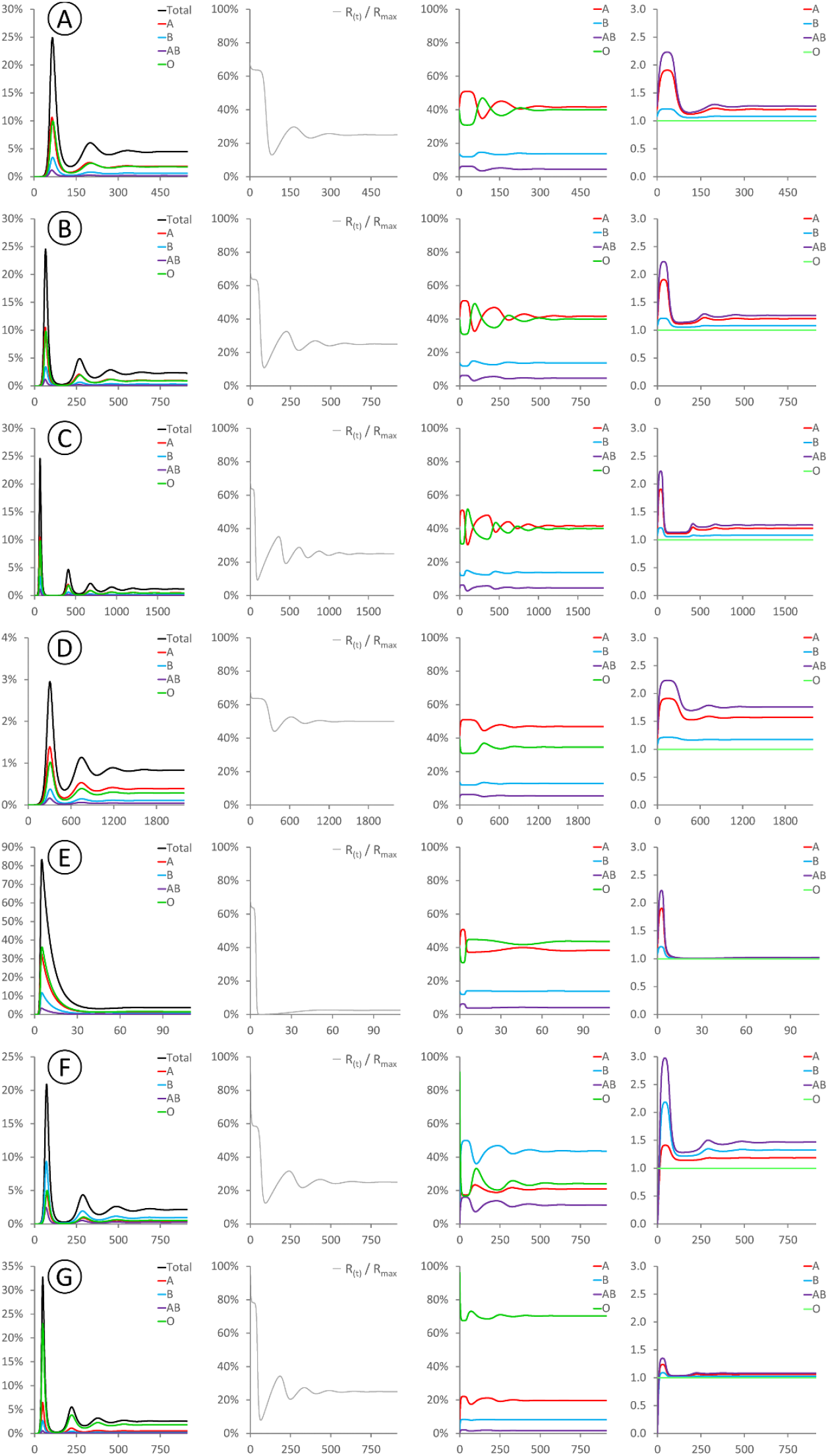
These panels show the evolution of various modelled parameters for epidemics involving waning immunity. Waning immunity is described by a parameter *ω* representing the rate of loss of immunity (see Methods for details). All epidemics were seeded at with type O index cases, and thus R(t) = R_max_ at time t=0. For all sub-panels, X axis denotes days. Y axes: (first column) proportion of the population infected; (second column) R(t) as a fraction of R_max_; (third column) distribution of blood types among currently infected individuals; (fourth column) cumulative risk of infection for each blood type relative to type O. Rows **A-E** are initialised with the same typical Western population blood type distribution used in Figures 1 and 2. **A:** R_max_ = 4, *ρ* = 20%, *ω*= 1/90 days^-1^. **B:** R_max_ = 4, *ρ* = 20%, *ω*= 1/180 days^-1^. **C:** R_max_ = 4, *ρ* = 20%, *ω*= 1/360 days^-1^. **D:** R_max_ = 2, *ρ* = 20%, *ω*= 1/90 days^-1^. **E:** R_max_ = 40, *ρ* = 20%, *ω*= 1/90 days^-1^. Row **F:** R_max_ = 4, *ρ* = 20%, *ω*= 1/90 days^-1^, initialised with an Indian population blood type distribution of 21.4%/39.9%/9.4%/29.3% type A/B/AB/O [17]. Row **G:** R_max_ = 4, *ρ* = 20%, *ω*= 1/90 days^-1^, initialised with a Peruvian population blood type distribution of 18.9%/8.1%/1.6%/71.4% type A/B/AB/O [18]. The relative risks to different blood groups during the final steady state are independent of *ω* (compare **A,B,C**), but depend on R_max_ (compare **B,D,E**) and on the background blood group distribution (compare **B,F,G**). The final case burden depends on all of R_max_, *ω, ρ* and the background blood group distribution.

Intriguingly, while the initial quasi-steady state is independent of R_max_ (see **Figure 2** above), the final steady state for an epidemic with waning immunity does depend on R_max_. When R_max_ is high, the final relative risk for all blood groups is close to unity. When R_max_ is low, then the final relative risk for all blood groups is similar to that seen in the initial quasi-steady state. Intuitively, this follows from the fact that for a highly contagious disease with a high R_max_, everyone will become infected as soon as their immunity wears off, irrespective of blood type. For a less contagious disease with a low R_max_, then there is more scope for differential susceptibility to play a part in how frequently individuals become infected.

## Discussion

### The biological interpretation of the parameter *ρ*

In this model, *ρ* represents the relative probability of virus transmission between an infected individual and an ABO-incompatible target individual. Mechanistically, this will encompass at least three sources of variability: (a) the extent to which the infected individual deposits ABO antigen on the surface of the virions produced, (b) the extent to which the target individual carries anti-A or anti-B antibodies, and (c) the ability of these antibodies to access the incoming virions and prevent infection.

Of these, (a) will depend not only on the host’s ABO genotype, but also on their Secretor (Se) status [21]. “Non-secretor” individuals are homozygous for null mutations in the FUT2 gene. In these individuals, ABO antigen is expressed exclusively in red blood cells and the vascular endothelium and is not present in other cell types. Conversely, in “secretor” individuals with at least one functional FUT2 allele, ABO antigens are found in virtually all cell types. In the context of the ABO-interference model described here, non-secretor individuals will still form anti-A and anti-B antibodies, and thus exhibit disease susceptibility according to their ABO blood group. However, they are unlikely to deposit A or B determinants on virions produced in lung cells, and so will transmit the virus freely, as if they were type O. Since around 20% of individuals in Western countries are non-secretors [22], and the estimated value of *ρ* is ∼35-40% (see above), then non-secretor individuals likely account for around half the observed cross-type transmission.

Factors (b) and (c) are related and I shall consider them together. While titres vary, virtually all individuals carry at least some antibodies to non-self ABO glycans. These are generally acquired in the first few years of life following exposure to microbial surface glycans similar to A or B determinants [12]. Anti-A and anti-B antibodies are typically IgM and IgA subtypes, however IgG can also be seen in patients following heterologous transfusion and systemic exposure to antigen. This has implications for factor (c) in that secretory IgA is the predominant antibody type found in mucosal secretions, along with smaller amounts of secreted IgM [23]. While the level of anti-ABO antibody in respiratory secretions has not been well studied, it is plausible that some amounts of both IgA and IgM may be present. These will not however be able to trigger complement-mediated inactivation of virus particles as the full complement cascade requires serum and is not present in the case of mucosal immunity. Anti-A/B antibodies may however block virus entry directly if the AB determinants are borne on the virus spike glycoprotein, as previously shown [10]. Alternatively, IgA and IgM may agglutinate virus particles and trap them in the mucus barrier layer.

### Implications of the estimated level of ABO-interference for public health strategy

If ABO-interference is the cause of the widely observed bias in SARS-CoV-2 infection rates among different blood types, then this models allows us to conclude that ABO incompatibility reduces SARS-CoV-2 transmission by at least 60% and potentially more. This implies that the apparent R0 for most of the largest epidemics around the world has already been suppressed by at least ∼25%, and that R_max_ is likely to be substantially higher than the actually-measured R_steady_. However, it is key to appreciate that no blood type is necessarily high- or low-risk: rather the nature of any protection is entirely context dependent. The presence of a diverse mix of blood types within any given community (i.e. within the pool of individuals that freely mix and may transmit the virus to each other) confers significant protection. Conversely, communities with limited blood type diversity have little or no inherent protection and will suffer disproportionately from SARS-CoV-2 infection. Herd immunisation threshold estimates derived from current data may therefore substantially underestimate the level of vaccination required to protect vulnerable communities such as Native populations in both North and South America. In these, the type O frequency approaches 100% [24], thus the true infectious potential of SARS-CoV-2 will be unmasked and the local R0 will tend towards R_max_.

This heterogeneity in transmissibility means that in general, the risk to non-O and in particular type AB individuals in most countries will be higher than risk to type O individuals, while type O individuals are more infectious than non-O individuals. This may contribute to the marked overdispersion in transmission frequency for SARS-CoV-2 [25], and help explain why a small subset of patients are responsible for the majority of transmission events. If other polymorphic surface glycans (e.g. Lewis and P antigens) behave similarly, this will further magnify the differences between “super-spreaders” and “super-recipients”.

Paradoxically, however, although in this model both disease vulnerability and infectiousness vary substantially between different blood types, it is important *<u>not</u>* to simplistically target vaccination on this basis. Rather, once a vaccine is available, care should be taken not to inadvertently destroy the existing blood type frequency structure that provides population-wise disease resistance, by ensuring good vaccine uptake among all communities. There is a danger that the growing public perception that “type O = low risk” will lead type O individuals to neglect or even refuse vaccination. If this tendency is not monitored and compensated for, it may disproportionately reduce the efficacy of public vaccination programs. Other types of blood-type-aware non-pharmaceutical interventions are not modelled here. If protective equipment is in limited supply, it may for example be appropriate for hospitals and care facilities to emphasise source control measures for type O “super-spreaders”, and recipient protection measures for type A and AB “super-recipients”. More sophisticated agent-based approaches will be needed to model this possibility [26]

### Implications of this model for evolution of the ABO polymorphism

Irrespective of the detailed epidemiology of any disease, at the individual level type O alleles are always selectively favoured under this model, while type A and B alleles are subject to frequency-dependent selection. This predicts that - as seen across the globe - type O will have the highest allele frequency, while A and B alleles will be at lower frequency and more nearly similar to each other. Extending the SIR model to cover the case of waning immunity shows that the elevated risk to non-O blood groups remains present even for endemic rather than epidemic disease, and thus long-term population morbidity and mortality from diseases subject to ABO-interference may be one factor affecting ABO allele population frequency. Evolutionarily, this model provides an interesting case where individual and group advantages differ, since whole O is always individually favoured, population-level disease resistance is optimised at relatively low type O frequencies. This tension between individual and group optima leads to a “tragedy of the commons” in which selection drives O alleles to a higher frequency than the group optimum and leaves the resulting population more vulnerable to disease.

### ABO-interference and seasonal coronaviruses with waning immunity

For seasonal coronaviruses, if these are subject to ABO-interference, this model predicts that that (in Western populations) type A and AB individuals should be more susceptible to infection, and type O less susceptible, but to a less pronounced degree than for SARS-CoV-2. It will therefore be interesting to determine whether non-O blood groups are indeed more likely to suffer more frequent repeat coronavirus infections. More speculatively, given the emerging evidence that COVID-19 is a multisystemic infection with particular impact on clotting pathways [27], could it be possible that the endemic coronaviruses may also have some effect on the likelihood of thrombosis? This could potentially explain the widely-observed phenomenon that type O individuals are on average slightly less likely to suffer from thrombosis and clotting-related myocardial infarction. Mechanistically, this is ascribed to an effect of ABO blood group on von Willebrand factor (vWF) levels, but the causal link by which ABO antigens regulate vWF levels remains obscure [28].

### Key observations and new experiments needed to test this model

In testing this model, the key experiment will be to directly determine whether A or B antigens are present on the virus envelope, and whether A- or B-specific antisera can neutralise virus from patients with the appropriate blood types. However, there are at least five other testable predictions from this model that may be addressable using existing epidemiological data:

i. in countries with high type B frequency such as India, type B individuals should be at higher risk than type A individuals.
ii. in studies of super-spreading events, the index cases should be disproportionately type O individuals and/or non-secretor individuals.
iii. Direct contact-tracing data should in general follow the blood transfusion rules. For in-family tracing, transmission between blood relatives - being more likely to share a blood type - may be more common than transmission between spouses, though this will be substantially confounded by age and behavioural effects.
iv. In hotspot areas, once the overall case frequency exceeds approximately 20%, the relative risks will begin to decline, as the epidemic proceeds through the type O population in a delayed manner.
v. In communities with high type O frequency, R_steady_ will be higher, the doubling time shorter and the relative risk to non-O blood types reduced when compared to otherwise similar communities with lower type O frequency. Similarly, communities with a highly skewed A:B ratio will have a higher R_steady_ and a shorter doubling time than communities with more nearly equal numbers of A and B individuals. This latter prediction has already been supported by two recent preprints [29, 30]

## Materials and Methods

### An SIR model of ABO-interference with virus transmission

A standard SIR model of infection divides the population into three compartments, <u>**S**</u>usceptible, <u>**I**</u>nfectious, <u>**R**</u>ecovered, representing the proportion of individuals in the population with each status, and thus S + I + R = 1. These compartments are linked by three differential equations:

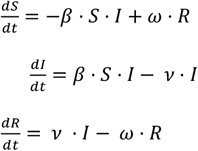

The parameters *β* and *ν* represent the rate constants for infection and recovery, and their reciprocals 1 / *β* and 1 / *ν* represent the average time required for one infectious individual to transmit to one susceptible individual, and the average duration of the infectious period. The product *β ·ν* represents the basic reproductive number R0. The parameter *ω* represents the rate constant for loss of immunity, and thus transfer from the R compartment back to S. Its reciprocal 1 / *ω* represents the average duration of immunity following recovery. In the analyses presented here, I extend this model by splitting each of S, I and R into four subcompartments representing the four ABO blood group phenotypes. These are then linked by a set of twelve equations:

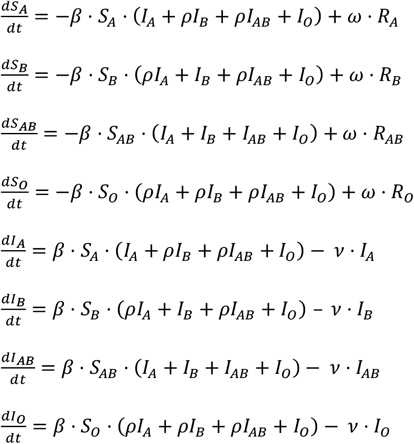

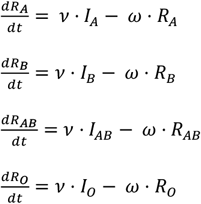

This can be graphically represented by the diagram on the right, with full arrows representing unimpeded transmission (infection rate = *β*) and dashed arrows representing impeded transmission (infection rate = *ρ ·β*).

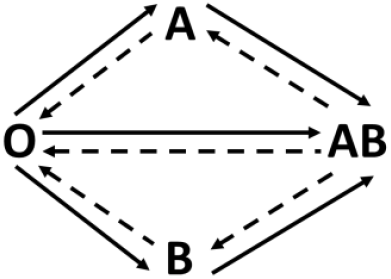

These equations were implemented as a Microsoft Excel spreadsheet (**Supplementary Data File 1**). In this model R0 is not a well-defined quantity since even in a fully susceptible population the current effective R value, denoted R(t), depends on *ρ* and on the blood type distributions in both S and I. In this paper, I use R_max_ to indicate the product *β ·ν*, i.e. the R0 that would be observed in the absence of any ABO-interference. This is the maximal possible value of R(t), and would be observed if *ρ* = 100%, or if all currently infected individuals are type O. For the work presented here, all epidemics were initiated by transferring 1/1,000,000 of the population from S to I at time t=0. This for example represents an initial importation of ∼9 infected index cases into a city the size of London. Varying these boundary conditions from 1/10,000 to 1/100,000,000 has no effect other than accelerating or retarding the initial progress of the epidemic (not shown). Except where otherwise specified in the main text, all index cases were assumed to be of blood group O. For all analyses except that presented in Figure 5, immunity was assumed to be permanent and thus *ω* = 0.

## Data Availability

All data and code required for this manuscript are contained within the text and supplements

## Acknowledgments

I would like to thank Dr Rupert Beale, Dr Jeremy Rossman, Dr Mike Clark and Mr C.F. Helix for helpful discussions. I am funded by the UK Higher Education Funding Council for England (HEFCE), the Leverhulme Trust (RPG-2019-414 194) and the Eastern Academic Research Consortium. I have no competing interests to declare. All data and code used in the analysis are available in the manuscript and supplementary materials.

## Supplementary Materials

**Data file S1:** Excel spreadsheet containing the implementation of the model described in the Methods.

